# Indian COVID-19 Preprints Submissions in bioRxiv and medRxiv Preprint Servers

**DOI:** 10.1101/2023.02.14.23285870

**Authors:** Narayanaswamy Vasantha Raju, Murtala Ismail Adakawa, N.S. Harinarayana, Chandrappa

## Abstract

This research aimed at determining the growth of deposition of preprints on servers by Indian researchers during COVID-19 pandemic. In the pre-pandemic period, the dominant server was arXiv upon which research from physics and other related domains have been the most predominant depositors. When the pandemic erupted and the need to share research findings became imperative, many previously dormant preprint servers received vibrant activations from several scientists across the globe. This is with the intention of bridging the gap between delays inherent in reviewing process and the dire need to share information for finding everlasting solutions to the raging pandemic. Many researchers, institutions, countries, etc. have contributed in this regard. The study used quantitative method and iSearch Portfolio expert-curated source for publications and preprints related to either COVID-19 or the novel coronavirus SARS-CoV-2 developed and maintained by National Institute of Health (NIH), US iSearch COVID-19 Portfolio. The study examined the Indian COVID-19 preprints deposited in bioRxiv and medRxiv preprint servers. The findings indicated that, Indian researchers have posted their papers in large numbers in bioRxiv and medRxiv servers with the medRxiv having the highest preprints (417, 40.44%) in 2020 against its counterpart bioRxiv (118, 10.96%) in the same year. Similarly, infectious diseases (except HIV/AIDS) (311) had the highest recurrence of the preprints submitted for deposition in servers. This is followed by epidemiology (263), public and global health (122), bioinformatics (59), among others. There is high collaboration among researchers who deposited their preprints in these servers where about 257 (24.93%) preprints were co-authored by 11+ authors followed by 3-authored and 4-authored with 124 preprints respectively (together accounts for 24.06%) and 2-authroed (114 (11.04%) preprints) respectively in a diminishing manner. The study concluded that, Indian researchers are actively participating in depositing preprints in servers notably bioRxiv and medRxiv.

## INTRODUCTION

When the immediacy runs into decision-making process concerning health emergency posed by COVID-19 pandemic, preprints serve as an intermediary between researchers, audience, and peer-review procedure (Majumder & Mandl, 2020; Otridge et al., 2022). Even though there has been yearning from scholars to change the nomenclature *preprints* to “*un-refereed manuscript”, “manuscript awaiting peer-review”, “non-reviewed manuscript*” (Ravinetto et al., 2021, p. 3), this is where a decision-support tool developed by Good Publication Practice (GPP), and International Committee of Medical Journal Editors (ICMJE) becomes imperative and relevant (Mathew et al., 2022). There is a differing perception on accepting preprint as a complete (Berg et al., 2016) or incomplete scientific manuscript (Añazco et al., 2021), which is still hanging and debatable. Completeness or incompleteness of preprints met COVID-19 that has set the stage for urgent need to discover and pressing need to share relevant information. The fact that, “*the potential benefits of preprints always outweigh the risks of harm*” (Ravinetto et al., 2021, p. 3), implies that, preprints provide evidence-based findings readily available for audience consumption. This is to the extent that, COVID-19 pandemic has changed research culture thereby generating, in a relatively short timeframe, a volume of research that even the most emergent fields such as deep learning or nanotechnologies have taken years to produce (Porter & Hook, 2020). This is mostly due to the sharing of research findings to public prior to publication thereby gaining maximum audience even if the journals are not open access that increases research productivity (Añazco et al., 2021). This follows from the urgency and imperative to share relevant information following the promulgation of the International Committee of Medical Journal Editor (ICMJE), WHO, and many scholarly journals urging authors to share their research findings to preprint servers prior to undergoing formal peer-review process (Añazco et al., 2021).

Acceleration of science and technology depends largely on the generation of new ideas fueled by creativity that provides innovations with a breathing possibility. One of the waysof creating new ideas isthe rapid production and dissemination of findings usually communicated through preprint servers or journals(Celi et al., 2021). As of June 25 2021, there were about 140,000 manuscripts on COVID-19 published or posted at PubMed, bioRxiv, and medRxiv (Tong et al., 2021), which indicates a new dawn of rapid evolution of research. Preprint server *arXiv* was the first server populated and popularized by physics, mathematics, and computer science community in 1991(Vlasschaert et al., 2020). Some researchers showed that, there have been preprints since 1961 but closed in 1967 due to resistance from journals (Otridge et al., 2022). To follow the queue, life sciences community also embraced preprint server *bioRxiv* founded in 2013 with over 75,000 preprints as of March 2020 (Vlasschaert et al., 2020). In 2018, from October to November alone, there were more than 2.2 million downloads wherein about 170 journals have collaborated with bioRxiv forming a process called B2J facilitating the transfer of preprints to journals for peer-review process (Vlasschaert et al., 2020). Unlike preprints in arXiv, researchers depositing their preprints in bioRxiv faced some criticisms as many journals flawed such attempts. However, many journals reviewed and reversed their policies and are now accepting preprinted papers (Vlasschaert et al., 2020).By June 2019, medRxiv preprint server was launched which aims to “*improve the openness and accessibility of scientific findings, enhance collaboration among researchers, document the provenance of ideas, and inform ongoing and planned research through more timely reporting of completed research*”(Vlasschaert et al., 2020, p. 2). Despite conference presentation of results, and educational blog postings, timely access to scientific findings contained on the preprints threatens the genuineness of investigations, as researchers show concern on the danger of using information contained on the preprints prior to undergoing peer-reviewed process (Vlasschaert et al., 2020). In spite of the observed challenges, preprints serve as an important ingredient for creativity and innovations. Perhaps this has relationship with rapid advancement in physics in developing equipment, perfecting methods of enquiry, improving measurement procedures especially in cosmological parameters used in radiation physics, nuclear forensics, radiotherapy, radiation oncology, etc.

This is true as Satish et al., (2020) have linked novelty of new ideas with change point detection analyzed via binary or bottom-up segmentation, availability of new terms and phrases in scientific publications, and time required for their appearance for public consumption. The rapidity of communicating scientific findings is visible through preprints despite containing hidden risk for public discourse (Celi et al., 2021)and that is why preprints continue to receive recognition for varying reasons. Some of the reasons include their 5 times citation rates against non-preprint scholarly outputs and their penetration power to reach scholarly community 14 months earlier than their counterpart non-preprint write-ups (Xie et al., 2021). Under traditional publishing condition, a manuscript takes at least 6 months of scrutiny by scientists (peers) (Fraser et al., 2021) using either blind, double, triple, etc. blind reviews depending upon the domain and quality of the manuscript. Among the proponents of peer review process, some agree that peer review process ensures rigor of the scientific findings while others opine that, it is slow, imperfect, and prone to partiality (Celi et al., 2021). This duality opened up windows for the introduction of preprint servers to serve as a remedy to the highlighted nagging challenges and an avenue through which scholars can share early-stage researches with high speed, open collaborations, and public scrutiny prior to submitting to journal for peer-review process (Celi et al., 2021). By so doing, scholars have discovered that, there is about 63 times increase in preprints distribution in the last 30 years but accounting for only 4% of the research articles published (Xie et al., 2021). There is observed shortening of number of days for publishing COVID-19 related articles over the non-COVID-19 submissions, which is within 120 days (Kodvanj et al., 2022).

## PREPRINTS AND ITS EVOLUTION

Preprints, disruptive force in scholarly communication, have become one of the major sources of scientific information with the potential of exponential growth and as a model of disseminating research findings (Vlasschaert et al., 2020). However, many researchers faulted the reliability of findings and cautioned authors and editors to“*check the accuracy of the citations and of the quotations of preprints before publishing manuscripts that cite them”*(Gehanno et al., 2022). To add to this argument, (Bero et al., 2021) compared the discrepancies of results in preprints and journal articles after publication and spin in interpretation. Out of the 67 preprints studied, 23(34%) had no discrepancy in preprints and journals, 15(22%) studies had at least one outcome mentioned in the journal not in preprint, and 8(12%) had one outcome mentioned in preprints only (Bero et al., 2021). Overall, they found that, results in preprints are largely similar to those reported in their corresponding journal, and cautioned that, reviewers should critically observe and evaluate discrepancies and spin in these research outputs (Bero et al., 2021). Probably, this is the reason why Kumar Verma et al., (2022) observed the hesitancy among health science librarians to take vaccine despite its development in 2021 due to the safety, negative information and confusion surrounding vaccine itself. This might have resulted due to the differences in attitudes towards depositing preprints in servers subscribed by the journal-specific domains. For instance, Yi and Huh, (2021) found that, out of 365 respondents in their research, 56 deposited submitted their manuscripts on preprint servers, with more than half had the attitude of preferring to deposit preprints, promote open access, get feedback on preprints, gain citations, etc. The researchers concluded that, there is a need for flexible policies for editors to accept preprints in Korea (Yi & Huh, 2021).

Preprints were common in arXiv server populated by physics, mathematics, computer science, but COVID-19 pandemic has sped up the rate at which scholars from other disciplines deposit their contents into other servers before review process takes place(Majumder & Mandl, 2020).This is true despite preprints are regarded as the *crude precursors of peer-reviewed papers*, they need proper scrutiny and assurance of stringent adherence to ethical policies prior to releasing them to audience for public consumption (Teixeira da Silva, 2021). Careful scrutiny of the high increase of preprints in the domain of biomedicine, COVID-19 pandemic has consolidated the rise of using these information resources to better understand the pattern of progressing of the virus and need to develop a vaccine (Majumder & Mandl, 2020).COVID-19 Science Update reported on topics ranging from health equity, vaccine, variants, natural history, testing, etc. (Otridge et al., 2022).

From the review, it is available in the literature that, policies surrounding preprint deposition are country-specific with some countries having high while others low deposition. For instance, Yi and Huh, (2021) conducted a research in Korea and found that, there is low usage of preprints in Korea despite some researchers showed positive attitudes towards depositing their researches in preprint servers. In other words, the research indicated that, researchers in engineering had prior knowledge on preprints than their counterpart medical researchers, which showed that discipline-specificity also plays a vital role in influencing preprints in preprint servers (Yi & Huh, 2021). As Indian research is still evolving, capturing these issues of significance is equally important in understanding the attitudes and behavior of researchers with respect to their willingness to deposit preprints in preprint servers. This will help policy formulators in a number of ways to implement relevant policies that could guide the conduct of meeting international standards in depositing preprints, reports, etc. prior to peer-review process. This implies the need to investigate the growth of preprint in a country wise or discipline wise to understand the pattern of contributions made by researchers in those countries. The current study is an attempt to understand the growth of preprints in India during the COVID-19 pandemic.

## RESEARCH QUESTION

The research attempts to provide answer to the following question

1. What is level of participation of Indian Authors in Depositing COVID-19 preprints in bioRxiv and medRxiv
2. What are the publication characteristics of Indian COVID-19 preprints, in terms of assessing the institutional contributions, identifying prolific authors, highly cited preprints and preprints with highest altmentric attention score (AAS) and understanding other publication characteristics

## METHODOLOGY

The data for the present study was obtained from iSearch COVID-19 Portfolio, a comprehensive, expert-curated source for publications and preprints related to COVID-19 or the novel coronavirus SARS-CoV-2, maintained by the National Institute of Health (NIH), US. This database indexes preprints deposited in seven preprint servers, including arXiv, bioRxiv, chemRxiv, medRxiv, preprints.org, Qeios, and Research Square, as well as peer-reviewed publications on COVID-19 indexed in PubMed.

For this study, we considered two major biomedical sciences preprint servers: bioRxiv and medRxiv. Out of the 44,594 preprints indexed in iSearch COVID-19 Portfolio on 29th November 2022, these two servers accounted for 25,429 preprints (14.70% for bioRxiv with 6555 preprints and 42.32% for medRxiv with 18874 preprints), which is 57.02% of the total preprints available in this database. The iSearch COVID-19 Portfolio was searched using the terms “2019-nCoV OR 2019nCoV OR COVID-19 OR SARS-CoV-2 OR Coronavirus AND India*” to retrieve the Indian preprints deposited in bioRxiv and medRxiv. Advanced search filters and search fields, such as Publication Date, Pub Types, Source filters, Publication (DOI, PMID), People (authors, author affiliation, first author, last author), and Content (title, abstract, full-text, condition, and supplemental text), were applied to obtain Indian COVID-19 preprints.

The preprints deposited from 01-01-2020 to 29-11-2022 were retrieved using the advanced search filter options of Publication Date, Pub Type as “preprints”, and Source as bioRxiv and medRxiv. This process resulted in obtaining 3,867 preprints, with 891 preprints found for bioRxiv and 2,970 preprints found for medRxiv using the keywords used in the study.

Initially, we used the “author location” option provided in the iSearch COVID-19 Portfolio database to limit the Indian COVID-19 preprints, but this process resulted in identifying only 143 preprints due to messy and blank entries in the downloaded Excel sheet. Hence, we abandoned this process and used Zotero open-source reference management software to obtain the PDF of the preprints through their DOIs. The PDFs of all 3,867 preprints were screened by two authors of this paper from December 1, 2022 to December 20, 2022 to identify Indian COVID-19 preprints in bioRxiv and medRxiv with at least one author associated with Indian institutions. This process resulted in obtaining 1031 Indian COVID-19 preprints, with 240 preprints deposited in bioRxiv and 791 preprints posted in medRxiv. This sample was used as the final data set for the study and further analysis. Figure 1 shows the flow chart of data collection process employed in the study.

**Figure 1:**
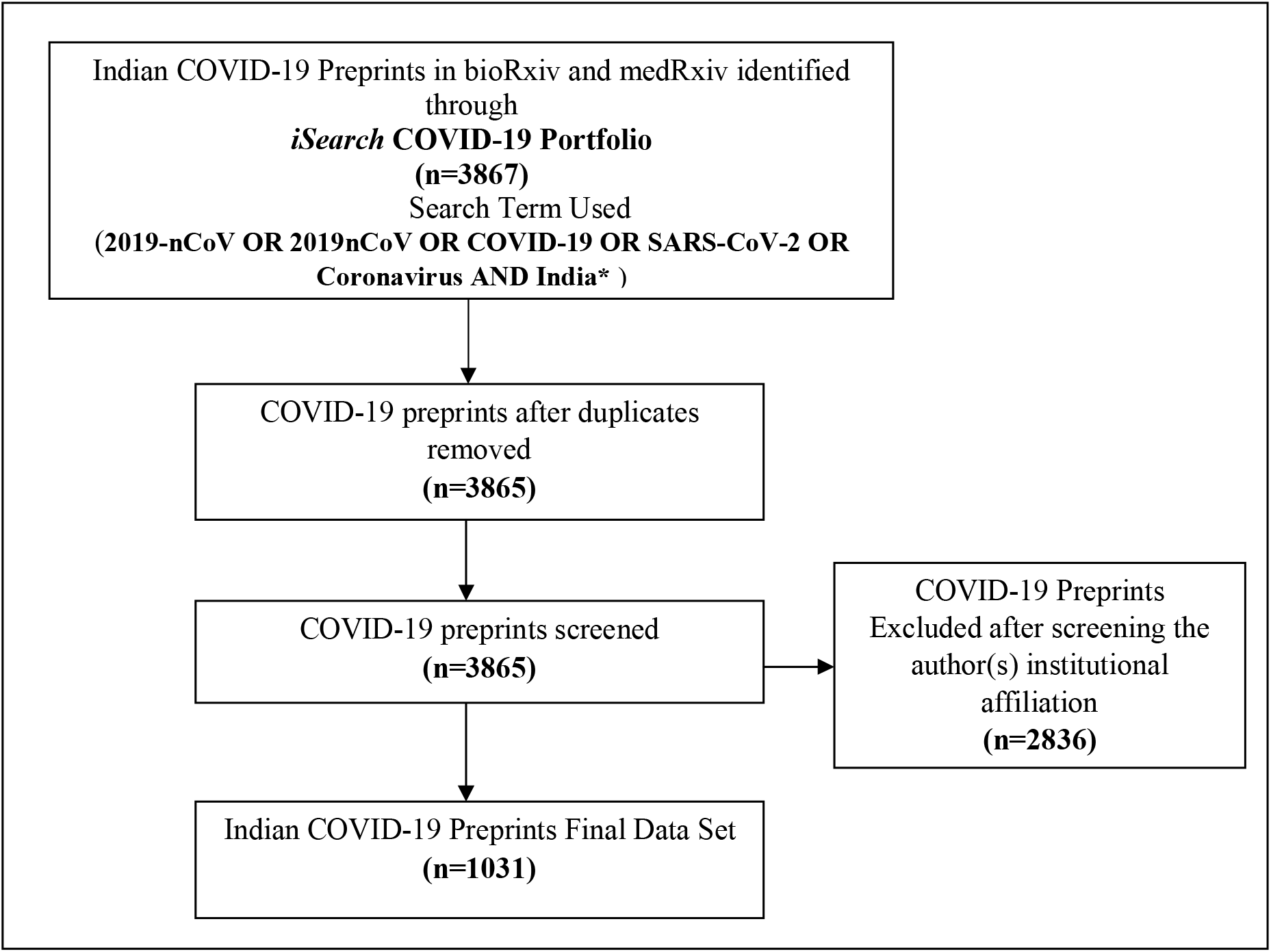
Flow Chart Depicting the Process of Data Collection.

To identify the Indian COVID-19 preprints that have received a high level of citations and have a high Altmetric Attention Score (AAS), we sourced our data from Dimensions (https://dimensions.ai), a vast and comprehensive scientific research database. In addition, we consulted the Retraction Watch (http://retractiondatabase.org/) database to uncover any reasons for retraction of the Indian COVID-19 preprints. The powerful reference management software, Zotero, was also utilized in this study. It effectively flags any retracted documents and provides easy access to these items through its item list feature.

## RESULTS

### 1. Indian COVID-19 Preprints in bioRxiv & medRxiv Preprint Servers

Figure 1 shows the year-wise distribution of Indian COVID-19 preprints (a) and total number of preprints posted in bioRxiv and medRxiv servers and total number of preprints by Indians (b). It is clear in the figure that, Indian researchers have contributed immensely in the scale up of preprints in the medRxiv (417, 40.44%) in 2020 against its counterpart bioRxiv (118, 10.96%) in the same year. Perhaps this resulted from the sudden upsurge of the COVID-19 pandemic and the rapid response researchers have undertaken in developing research-based interventions that could inform stakeholders on how to tackle the spread of the virus particle. Even though in the following year 2021, there was a little decline in the posting of preprints during this period owing to the development of vaccines and relative herd immunity, Indian researchers did not slacken in their quest to finding everlasting solutions to the pandemic. This resulted in generation of 272 (26.38%) preprints in medRxiv and 92 (8.9%) in bioRxiv in the year 2021.

The implication of this finding is that, Indian scholars have not been reluctant in the fight against the virus and contributed towards development of researches that aided in the vaccine development, logistics, to mention but a few. This agrees with submission of Singh et al., (2020) who suggested that, Indian scholars should deposit their researches in such repositories. In addition, this is not a surprise as Porter and Hook, (2020) have indicated that, COVID-19 has affected research production greater than deep learning and nanotechnology have produced combined. From another perspective, in the year 2022, the number of preprint deposition drastically reduced 102 (9.89%) in medRxiv and 35 (3.39%) in bioRxiv owing to probably the development of vaccines and reduction of the number of infections in the population. There are 791(76.72%) preprints deposited in medRxiv and 240 (23.28%) preprints in bioRxiv. Overall there are 1031 preprints submitted by the Indian authors from the beginning of 2020 to November 2022.

**Figure 1:**
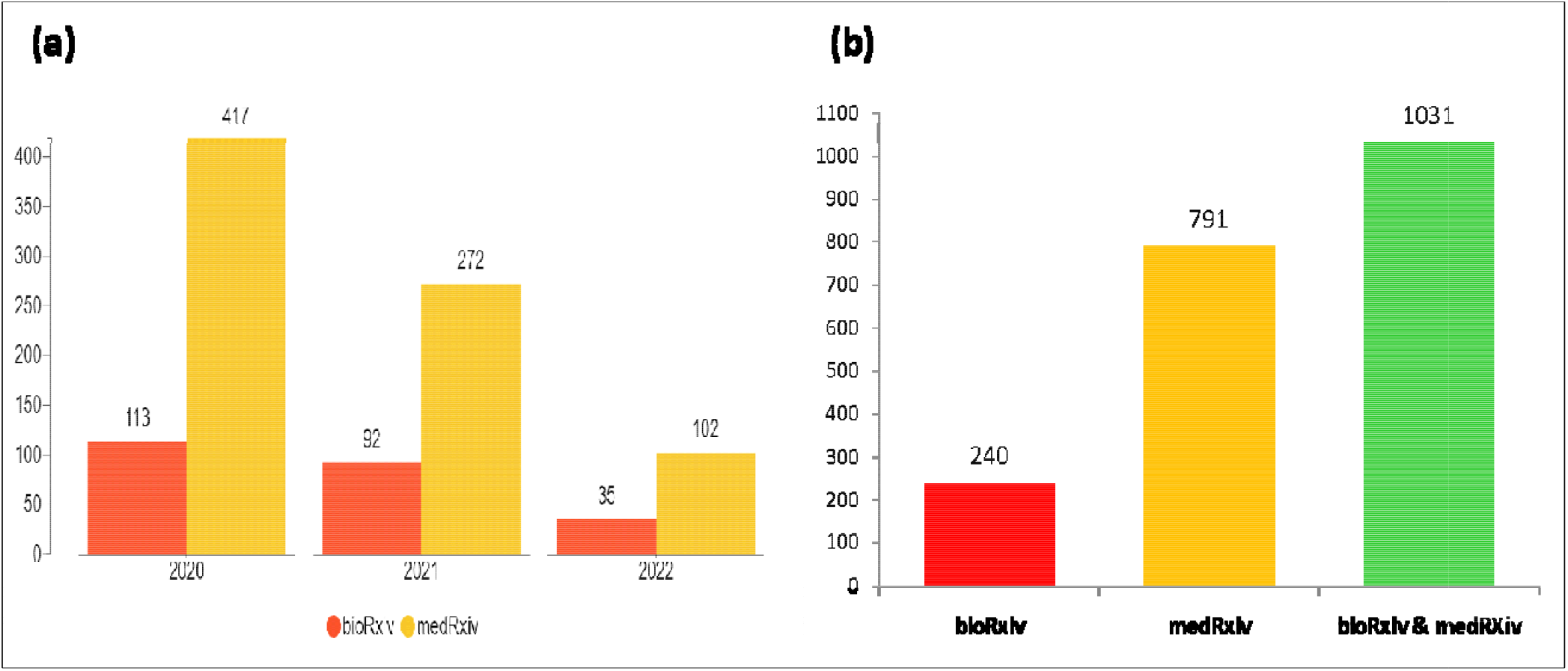
Year-Wise Distribution of Indian COVID-19 Preprints (a) and COVID-19 Preprints Submitted to bioRxiv and medRxiv Preprint Servers (b)

### 2. License Types of Indian COVID-19 Preprints

Figure 2 presents the preprint license distribution among researchers. The data shows that 42.58% of the researchers selected the CC BY-NC-ND 4.0 license, followed by 32.30% choosing All Rights Reserved, 10.67% choosing CC BY-ND 4.0, 9.12% choosing CC BY 4.0, 4.85% choosing CC BY-NC 4.0, and only 0.48% choosing CC0. This aligns with the findings of Fraser et al., (2021) who reported that authors have the option to pick from several Creative Commons licenses when uploading their preprints to bioRxiv and medRxiv servers. The Creative Commons licenses include CC0 (No Rights Reserved), CC BY 4.0 (Attribution), CC BY-NC 4.0 (Attribution, Non-commercial), CC BY-ND 4.0 (Attribution, No Derivatives), and CC BY-NC-ND 4.0 (Attribution, Non-commercial, No Derivatives). A large portion of Indian researchers, 42.58%, selected the CC BY-NC-ND 4.0 license, which allows for the sharing of knowledge without any limitations for public use.

**Figure 2:**
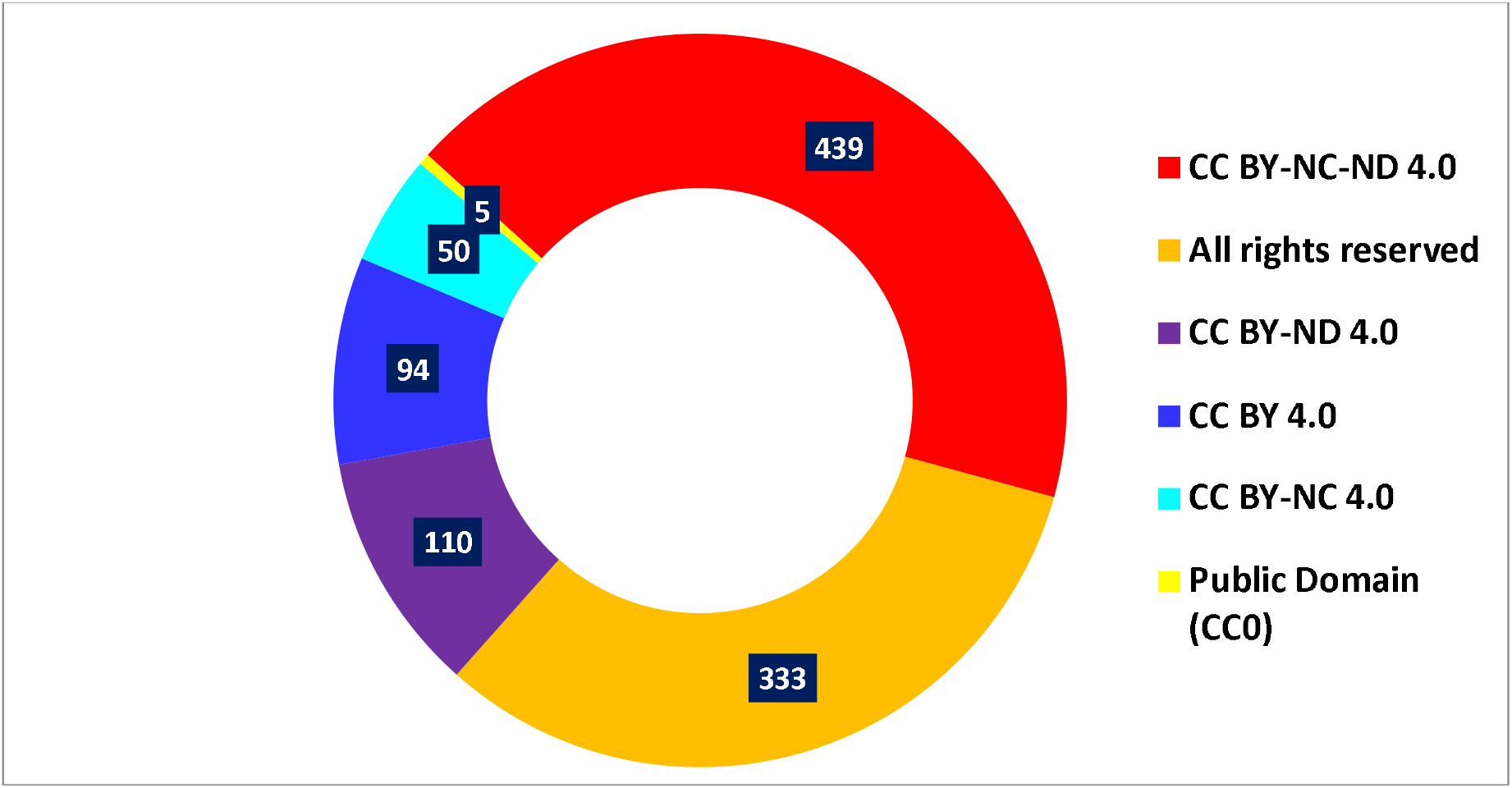
Preprint License Type.

### 3. Subject Categories in Which 10 or More COVID-19 Preprints Published

Figure 3 illustrates the distribution of contributions in different subject categories made by Indian scholars. The data reveals that the most frequently submitted preprints in servers were related to infectious diseases (other than HIV/AIDS) with 27.74% (286 preprints). Epidemiology came in second with 22.89% (236 preprints), followed by public and global health (9.89%, 102 preprints) and bioinformatics (5.91%, 61 preprints). These findings align with the study conducted by Fraser et al., (2021) which found that the subject areas of preprints deposited in servers were not limited to biomedical research only. This suggests that the COVID-19 pandemic has encouraged interdisciplinary collaboration among researchers, leading to the examination of the challenges posed by the pandemic from multiple perspectives (Porter & Hook, 2020).

**Figure 3:**
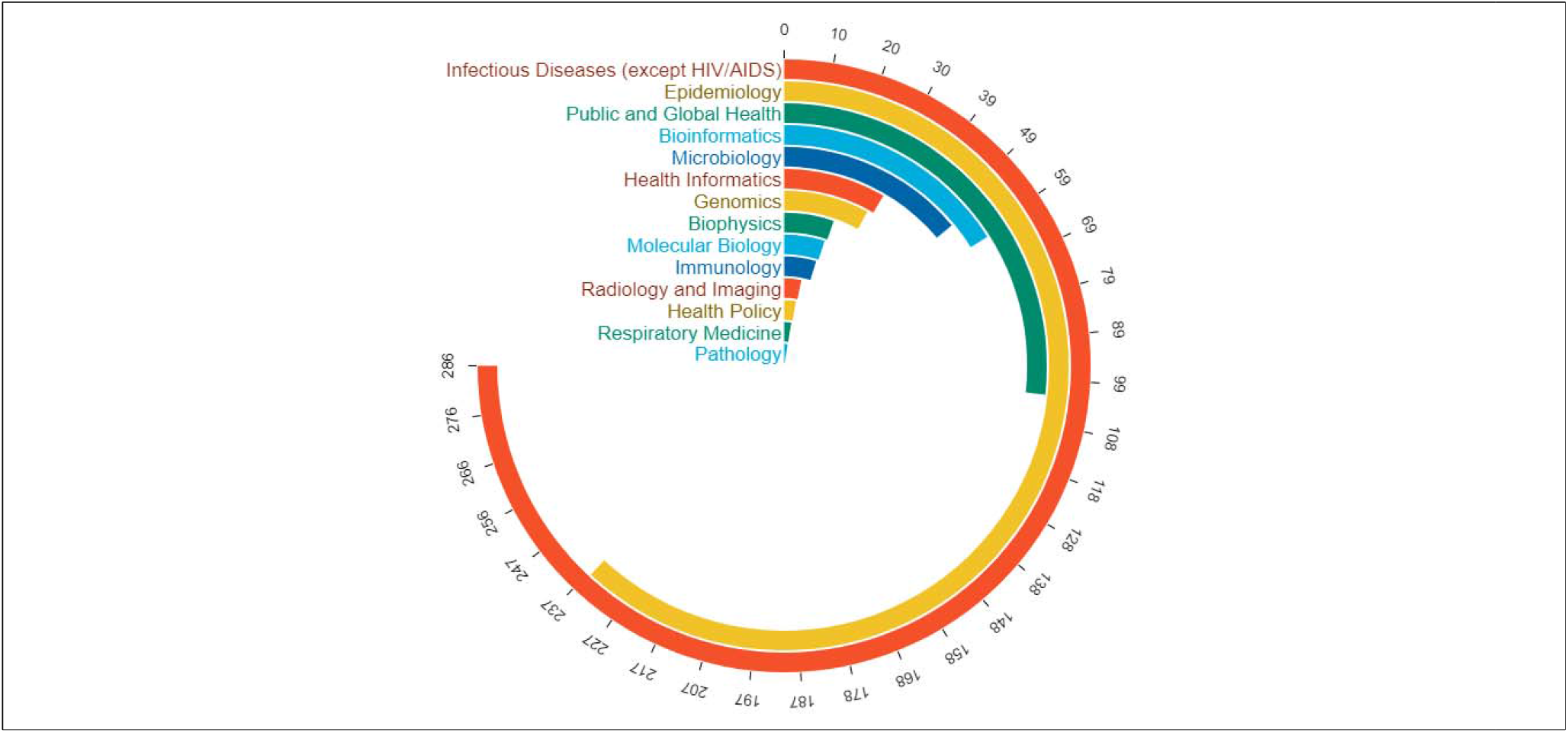
Top Indian COVID-19 Preprints Subject Categories.

The co-occurrence keyword network visualization was produced using the VOSviewer bibliometric network software (version 1.6.18). The network diagram was created by using keywords that appeared 6 or more times, out of the 4047 terms. A total of 163 terms met the threshold and resulted in the formation of five major clusters, as displayed in Figure 3(a). Cluster 1 (in red) consists of 28 keywords with terms such as “mathematical model”, “forecasting”, “country”, “peak”, “trend”, “number”, “modeling”, and others. Cluster 2 (in green) consists of 24 keywords with terms mainly related to “hospital”, “severe covid”, “vaccination”, and “Indian population”, among others. Cluster 3 (in blue) has 23 keywords with terms such as “sars-cov”, “variant”, “mutation”, and “detection”. Cluster 4 (in yellow) has 12 keywords including “vaccine”, “healthcare workers”, “chadox1 ncov”, “Kerala” and “Tamil Nadu”, and others. Cluster 5 (in purple) contains 11 keywords with terms like “safety”, “vaccine efficacy”, “bbv152”, “evaluation” and others. These clusters highlight the significant concentration of COVID-19 studies conducted in India

**Figure 3(a):**
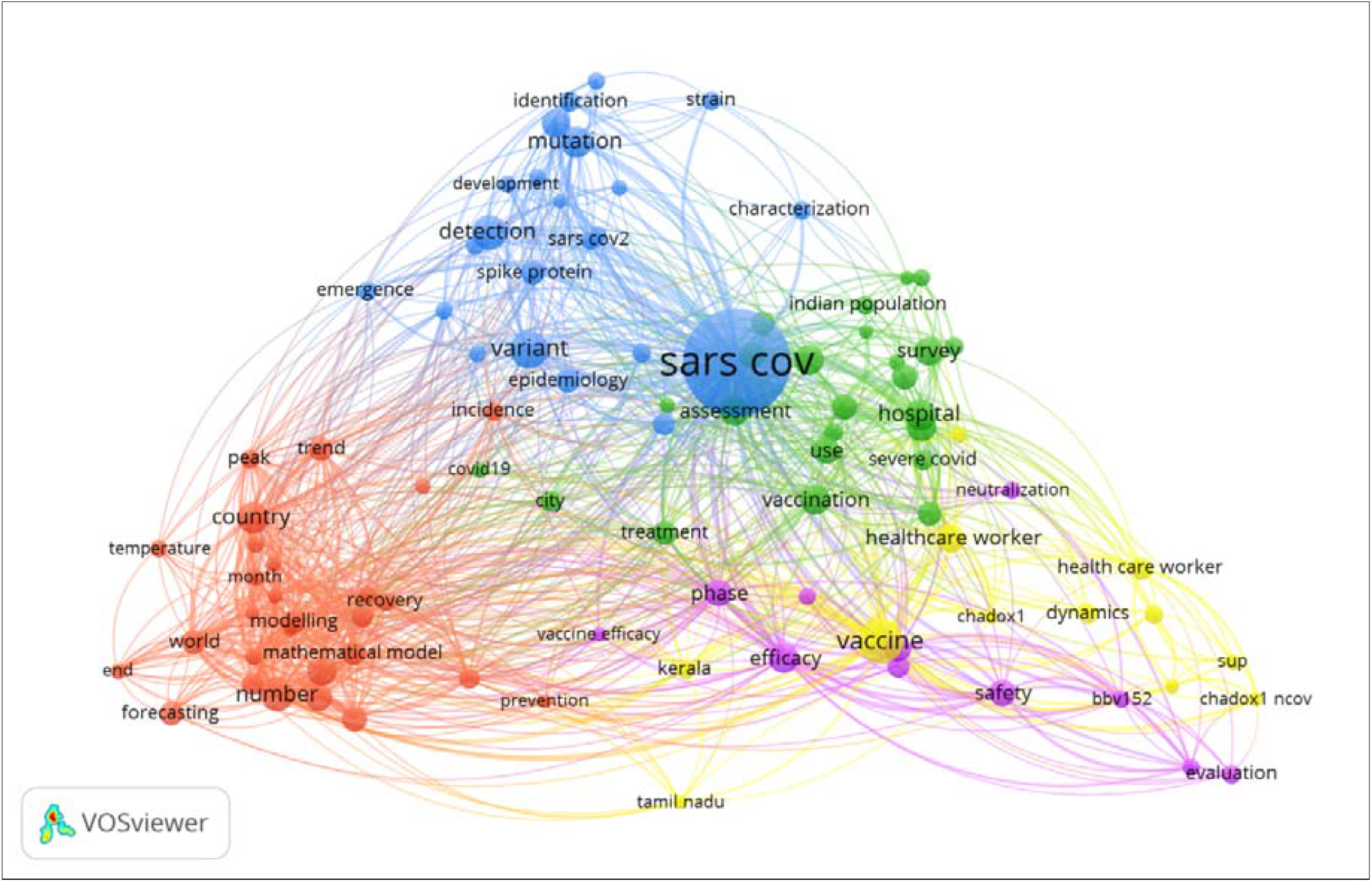
Co-occurrence of Keywords in Indian COVID-19 Preprints.

### 4. Number of Authors Associated with COVID-19 Preprints

Figure 4 show the number of authors in a publication. From the figure, it is observable that, about 257 (24.93%) preprints were co-authored by 11+ authors. This is followed by 3-authored and 4-authored with 124 preprints respectively (together accounts for 24.06%) and 2-authroed (114 (11.04%) preprints) respectively in a diminishing manner. This implies that, there is rise of collaboration among Indian scholars due to the pandemic and this pattern of collaboration has helped in finding suitable solutions to the upsurge of the pandemic thereby developing vaccines. As Porter and Hook, (2020) reiterated that, when a field develops rapidly, as in the case with COVID-19, there is observable change in the research community, which includes new behavior, change in collaboration pattern, and uses of information collectively shape the field. This agrees with Waltman, et al., (2021, p. 73) who called for collaboration for inter-sectoral and international agencies, and recommended for collaboration among funders, governmental organizations, research institutions, and publishers to align their data “around a principle “as open as possible and as closed as necessary”. These collaborations, geared towards preprinting and data sharing must be in a systematic and sustained way accompanied by monitoring and accountability mechanisms (Waltman, et al., (2021). From another perspective, these collaborations led to same-subject collaboration, same-institutional collaboration, increased in biological sciences and medicine collaborations, and no significant new research relationship pairings outside medicine/biology (Porter & Hook, 2020). This means that most collaborations are in-field with respect to medicine and biological sciences.

**Figure 4:**
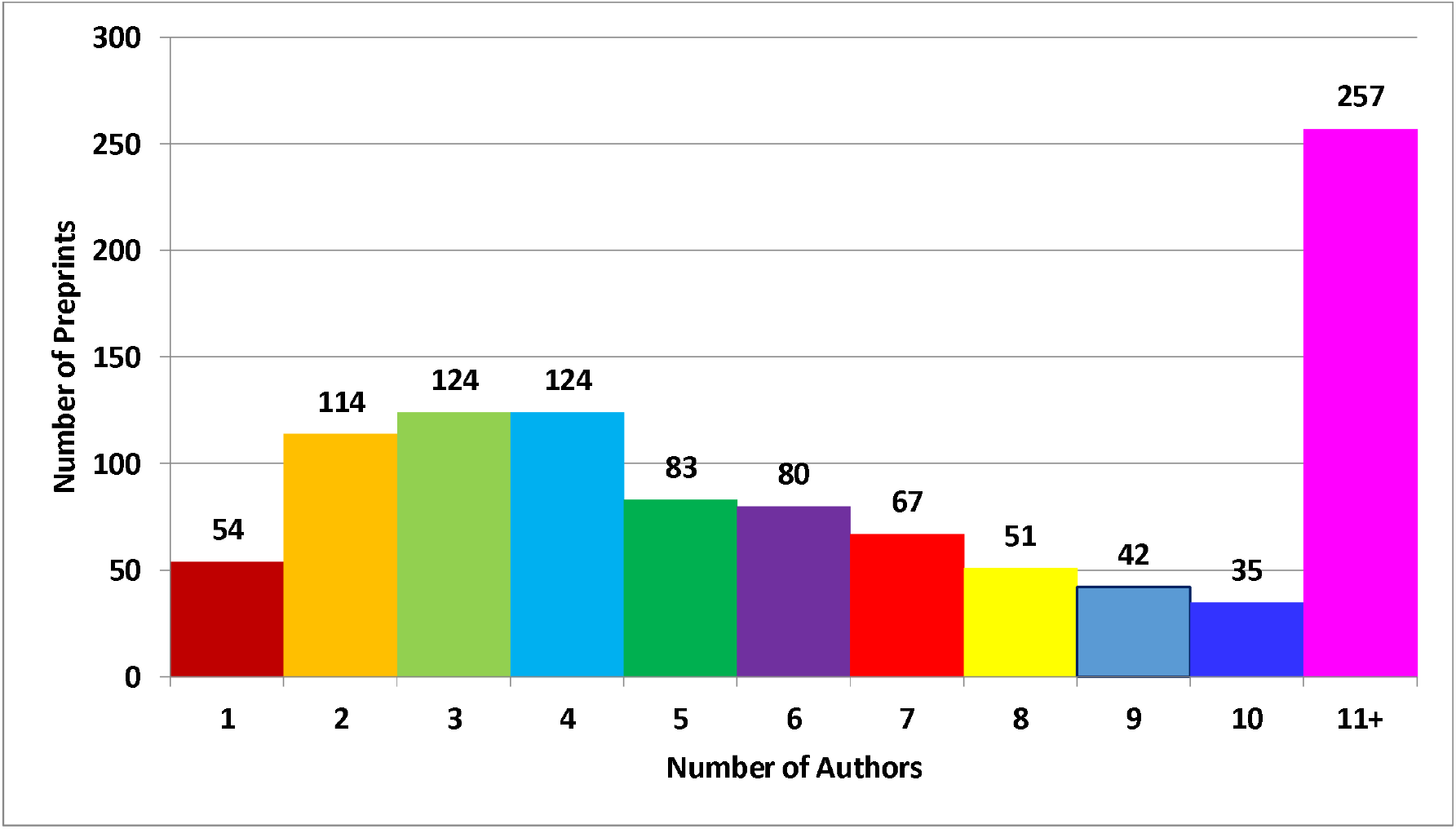
Number of Authors Associated with Each COVID-19 Preprints.

### 5. Prolific Authors and Institutions with more than 10 COVID-19 Preprint

The figure 5 showcases the top authors (a) and institutions (b) that have published 10 or more COVID-19 preprints in India. The table reveals that Priya Abraham had the most preprints, with 16 accounting for 1.55% of the authors with more than 10 preprints in India. Other top authors include Madhvi Joshi and Samiran Panda with 14 preprints each, and Balram Bhargava and Nivedita Gupta with 13 preprints each. The data indicates that Indian researchers are actively contributing by depositing preprints in servers. Most of the top authors are affiliated with the ICMR and its institutions and have published preprints collaboratively. There is also a pattern where authors affiliated with their own institutions collaborate more often than with external authors or institutions. The co-authorship network analysis further highlights the connections between the top authors from ICMR. Priya Abraham of ICMR-NIV, Pune has the strongest connections, with 113 links, and her main collaborators are Samiran Panda, Balram Bhargava, Nivedita Gupta, Pragya Yadav, Deepak Y Patil, and others. (<u>Supplementary File-1</u> for main collaborator of Priya Abraham (Figure-S1) derived from VOSviewer network visualization application version 1.6.18)

In Figure 5 (b), the top Indian institutions with the highest COVID-19 preprints are listed. The AIIMS in New Delhi has the most preprints with 43, followed by the IISc in Bengaluru with 26 preprints and the ICMR-NIV in Pune, with 22 preprints. The other top institutions, as shown in Figure 5(b), mainly work in the medical and biomedical sciences field, with the exception of the IIPS (International Institute of Population Sciences), which conducted studies related to mathematical models and infectious diseases.

**Figure 1:**
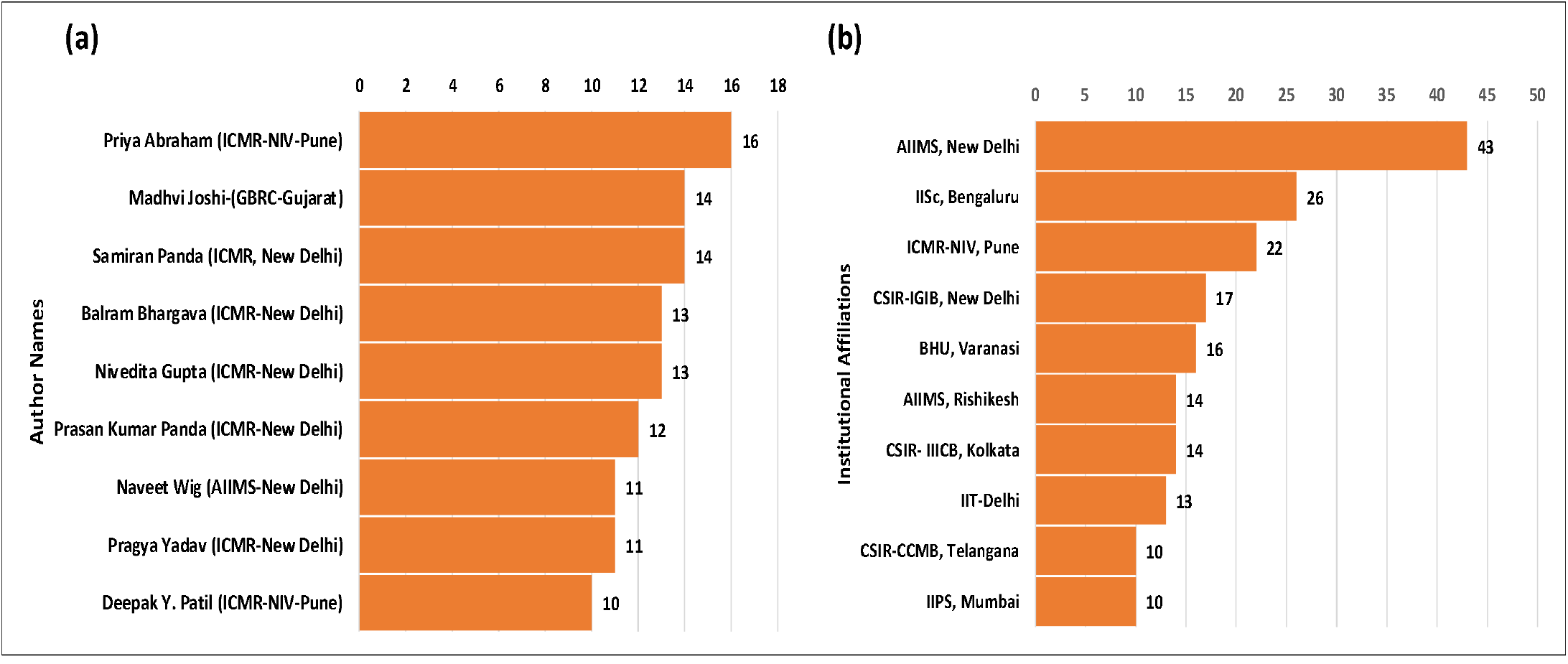
Authors (a) and Institutions (b) with More than 10 COVID-19 Preprints.

### 6. COVID-19 Preprints Published in Journals

Figure 6 presents a clear illustration of the number of COVID-19 related preprints published in scholarly journals. The figure depicts that a total of 1031 preprints were submitted to preprint servers, with 240 of them deposited in bioRxiv and 791 submitted to medRxiv. Out of the 240 preprints submitted to bioRxiv, 118 (49.17%) were eventually published in scholarly journals. On the other hand, 165 (20.86%) of the 791 preprints submitted to medRxiv were published in journals. This data adds up to a total of 283 (27.45%) preprints published in scholarly journals.

**Figure 6:**
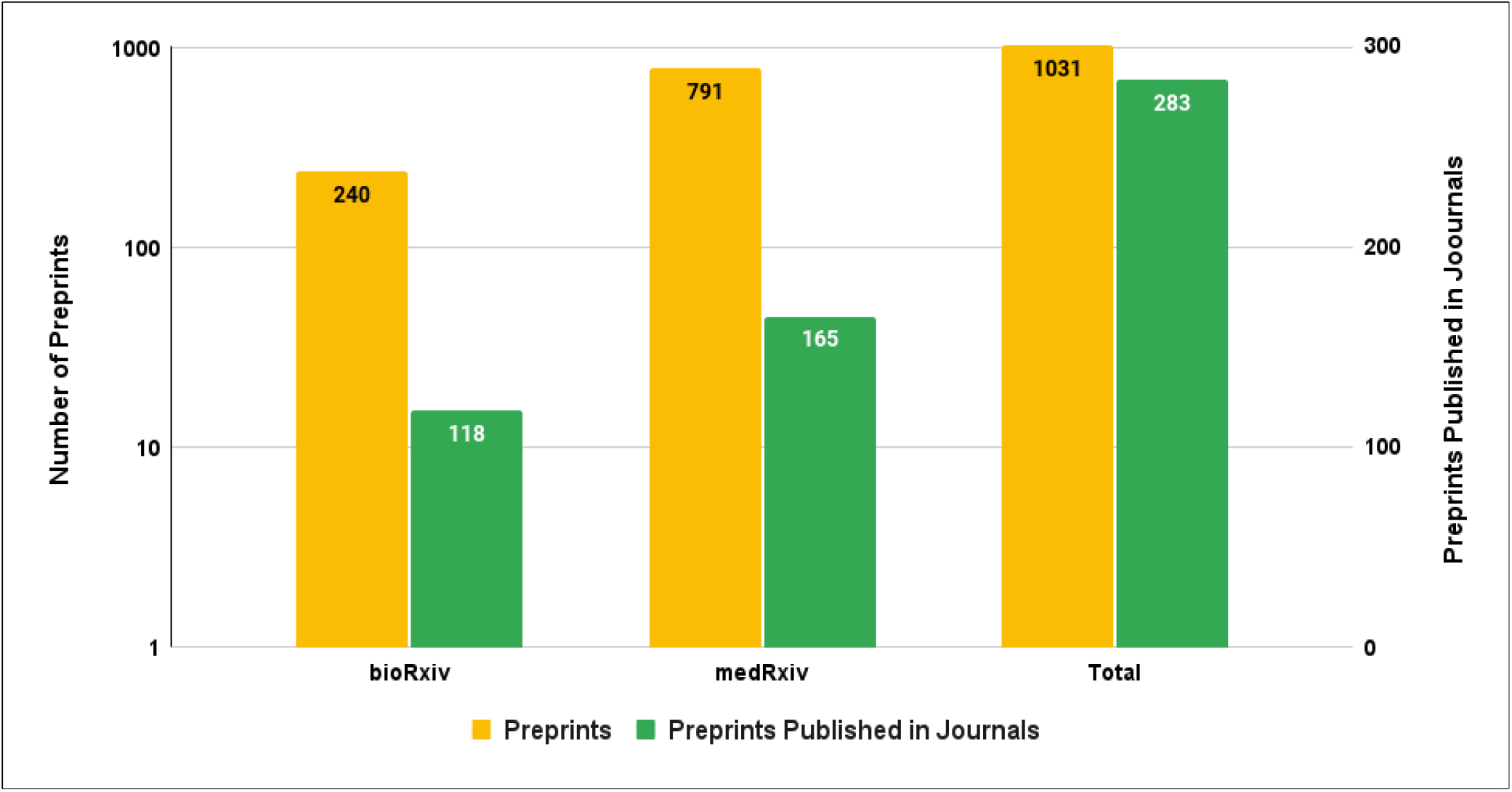
Number of COVID-19 Preprints and Their Appearance in Journals.

The figure also highlights that almost half of the preprints submitted to bioRxiv were published in scholarly journals. This suggests a shift in the way research is communicated, with researchers increasingly choosing to deposit their preliminary results in preprint servers rather than submitting them directly to journals. This shift highlights the expanding scope of research communication through the use of preprint servers in the current era.

### 7. Top Journals Which Published Indian COVID-19 Preprints

Table 1 showcases the journals that have published 3 or more Indian COVID-19 preprints. The top two journals in this category are PLOS ONE, a renowned multidisciplinary journal, and Scientific Report, another multidisciplinary journal. PLOS ONE has published 14 Indian authored preprints (Impact Factor: 3.752%) and Scientific Report has published 10 preprints (Impact Factor: 4.997%). These results show that the high quality of Indian research is being recognized globally, as these preprints are being accepted and published by top impact factor and high quartile ranking journals. The B2J and M2J options available in bioRxiv and medRxiv allow authors to easily transfer their manuscript to journals without resubmitting or formatting it. Many major publishers, including PLOS, are now encouraging authors to deposit their preprints before submitting them to journals. This further reinforces the idea that preprint servers are an effective and efficient way to communicate research results.

**Table 1:**
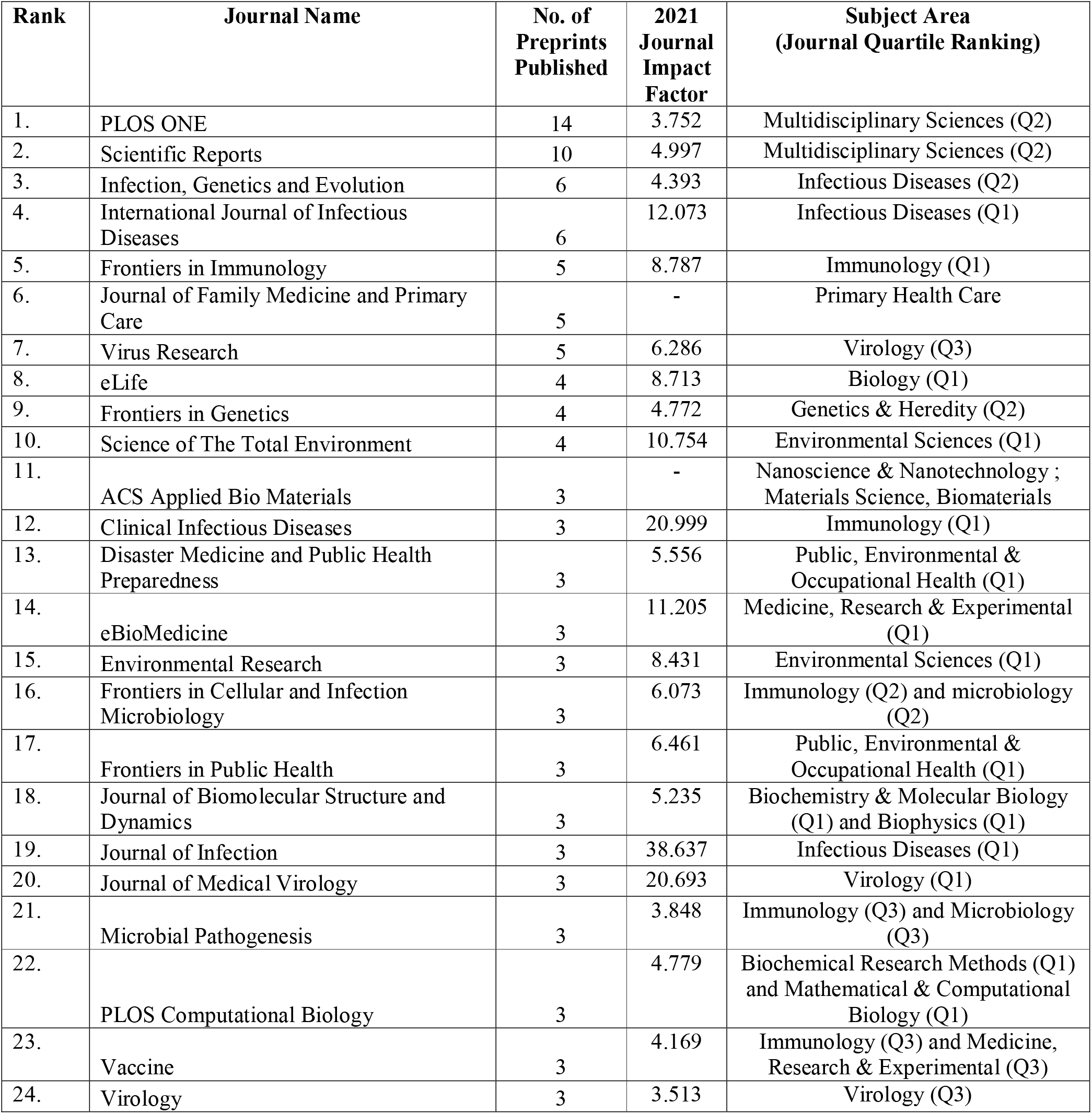
Journals Published 3 or More Indian COVID-19 Preprints.

### 8. Highly Cited Indian COVID-19 Preprints

Table 2 showcases the Indian COVID-19 preprints with high citations. It is evident that both bioRxiv and medRxiv, the servers that house these research outputs, are highly sought after. The preprint “Repurposed antiviral drugs for COVID-19-interim WHO solidarity trial results” is the most cited with 263 citations. This preprint was in high demand due to its crucial role in determining effective pharmaceutical treatments for COVID-19 patients. The drugs featured in this study, remdesivir and hydroxychloroquine, received a lot of attention, however, the results showed that these repurposed antiviral drugs had little to no impact on hospitalized COVID-19 patients (WHO Solidarity trial consortium et al., 2020). Following this preprint, “Convergent evolution of SARS-COV-2 spike mutations, L452R, E484Q, and P681R, in the second wave of COVID-19 in Maharashtra, India” has 227 citations and “SEIR and regression model-based COVID-19 outbreak predictions in India” has 179 citations, coming in third place. Table 3 showcases other highly cited preprints. These preprints are primarily focused on subjects such as genomics/molecular biology, bioengineering/radiology and imaging, and health informatics (predictive models based on machine learning).

**Table 2:**
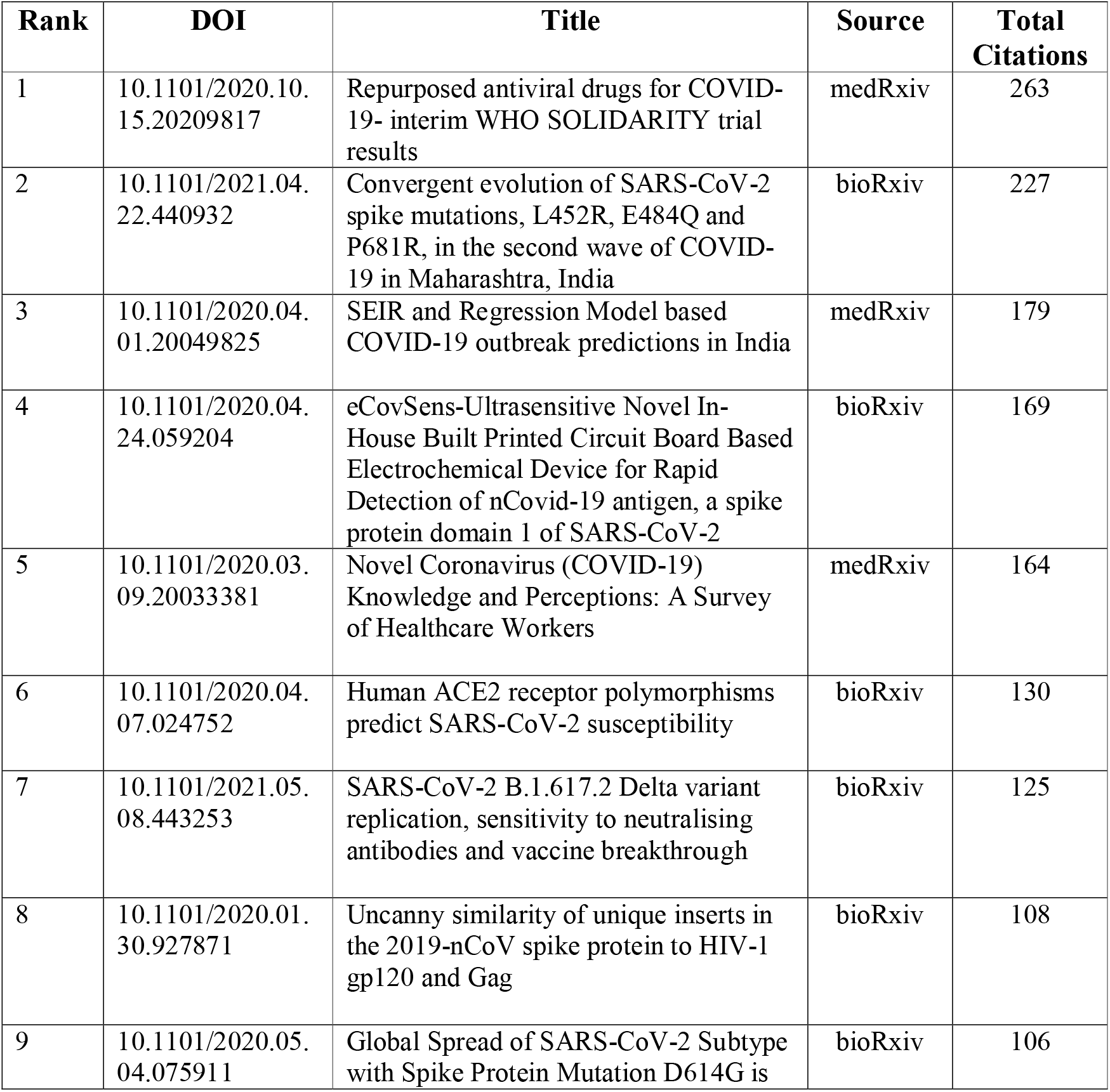
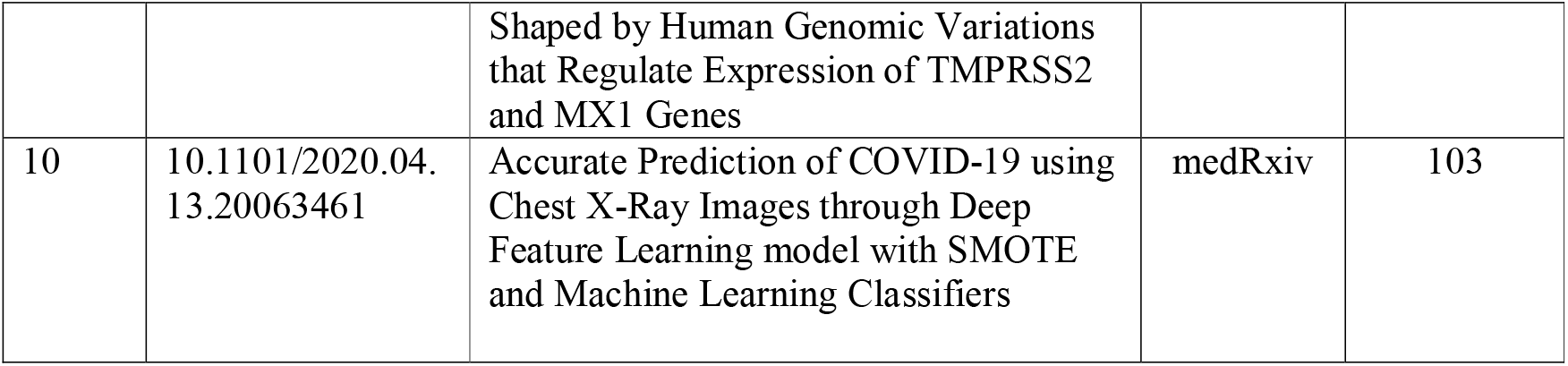
Highly Cited Indian COVID-19 Preprints.

| Rank | DOI | Title | Source | Total Citations |
| --- | --- | --- | --- | --- |
| 1 | 10.1101/2020.10.15.20209817 | Repurposed antiviral drugs for COVID-19- interim WHO SOLIDARITY trial results | medRxiv | 263 |
| 2 | 10.1101/2021.04.22.440932 | Convergent evolution of SARS-CoV-2 spike mutations, L452R, E484Q and P681R, in the second wave of COVID-19 in Maharashtra, India | bioRxiv | 227 |
| 3 | 10.1101/2020.04.01.20049825 | SEIR and Regression Model based COVID-19 outbreak predictions in India | medRxiv | 179 |
| 4 | 10.1101/2020.04.24.059204 | eCovSens-Ultrasensitive Novel In-House Built Printed Circuit Board Based Electrochemical Device for Rapid Detection of nCovid-19 antigen, a spike protein domain 1 of SARS-CoV-2 | bioRxiv | 169 |
| 5 | 10.1101/2020.03.09.20033381 | Novel Coronavirus (COVID-19) Knowledge and Perceptions: A Survey of Healthcare Workers | medRxiv | 164 |
| 6 | 10.1101/2020.04.07.024752 | Human ACE2 receptor polymorphisms predict SARS-CoV-2 susceptibility | bioRxiv | 130 |
| 7 | 10.1101/2021.05.08.443253 | SARS-CoV-2 B.1.617.2 Delta variant replication, sensitivity to neutralising antibodies and vaccine breakthrough | bioRxiv | 125 |
| 8 | 10.1101/2020.01.30.927871 | Uncanny similarity of unique inserts in the 2019-nCoV spike protein to HIV-1 gp120 and Gag | bioRxiv | 108 |
| 9 | 10.1101/2020.05.04.075911 | Global Spread of SARS-CoV-2 Subtype with Spike Protein Mutation D614G is | bioRxiv | 106 |
|  |  | Shaped by Human Genomic Variations that Regulate Expression of TMPRSS2 and MX1 Genes |  |  |
| 10 | 10.1101/2020.04.13.20063461 | Accurate Prediction of COVID-19 using Chest X-Ray Images through Deep Feature Learning model with SMOTE and Machine Learning Classifiers | medRxiv | 103 |

**Table 3:** COVID-19 Preprints with Highest Altmetrics Attention Score (AAS)

| Rank | DOI | Title | Source | AAS |
| --- | --- | --- | --- | --- |
| 1 | 10.1101/2020.01.30.927871 | Uncanny similarity of unique inserts in the 2019-nCoV spike protein to HIV-1 gp120 and Gag | bioRxiv | 13637 |
| 2 | 10.1101/2020.10.15.20209817 | Repurposed antiviral drugs for COVID-19- interim WHO SOLIDARITY trial results | medRxiv | 8068 |
| 3 | 10.1101/2021.08.20.21262342 | Emergence and phenotypic characterization of C.1.2, a globally detected lineage that rapidly accumulated mutations of concern | medRxiv | 4133 |
| 4 | 10.1101/2021.06.30.21259439 | Efficacy, safety, and lot to lot immunogenicity of an inactivated SARS-CoV-2 vaccine (BBV152): a, double-blind, randomised, controlled phase 3 trial | medRxiv | 2924 |
| 5 | 10.1101/2022.08.23.504936 | SARS-CoV-2 Omicron spike mediated immune escape and tropism shift | bioRxiv | 2357 |
| 6 | 10.1101/2021.04.23.441101 | Neutralization of variant under investigation B.1.617 with sera of BBV152 vaccinees | bioRxiv | 2292 |
| 7 | 10.1101/2021.05.08.443253 | SARS-CoV-2 B.1.617.2 Delta variant replication, sensitivity to neutralising antibodies and vaccine breakthrough | bioRxiv | 2008 |
| 8 | 10.1101/2021.07.23.21260716 | COVID-19 Associated Hepatitis in Children (CAH-C) during the second wave of SARS-CoV-2 infections in Central India: Is it a complication or transient phenomenon | medRxiv | 2005 |
| 9 | 10.1101/2020.09.03.20187252 | Convalescent plasma in the management of moderate COVID-19 in India: An open-label parallel-arm phase II multicentre randomized controlled trial (PLACID Trial) | medRxiv | 1851 |
| 10 | 10.1101/2022.01.04.21268536 | Prevalence, characteristics, and predictors of Long COVID among diagnosed cases of COVID-19 | medRxiv | 1759 |

### 9. Top 10 Indian COVID-19 Preprints with Highest Altmetrics Attention Score (AAS)

Table 3 reveals the most buzz-generating COVID-19 preprints, as indicated by their Altmetric Attention Scores (AAS). The AAS measures the attention an article receives from various sources, including media outlets, policy documents, blogs, social media, and more. Determined through dimenions.ai, which tracks and indexes altmetric data for each article along with citations, the AAS provides a glimpse into the reach and influence of a given publication.

Topping the list with an AAS of 13637 is the preprint “*Uncanny similarity of unique inserts in the 2019-nCoV spike protein to HIV-1 gp120 and Gag*”. When this preprint was first published on bioRxiv in January 2020, it sparked a heated debate on social media over its claim that COVID-19 was man-made. However, the preprint was soon retracted due to a lack of substantial evidence to support its claims. Next on the list is the preprint “*Repurposed antiviral drugs for COVID-19-interim WHO SOLIDARITY trial results*”, with an AAS of 8068. With the world anxiously searching for effective treatments for COVID-19 during the pandemic”s first wave, this preprint, which examined the efficacy of existing antiviral drugs like remdesivir, hydroxychloroquine, lopinavir, and interferon, received a lot of global attention. The transnational study “*Emergence and phenotypic characterization of C*.*1*.*2, a globally detected lineage that rapidly accumulated mutations of concern*” has received AAS of 4133. Other preprints which have received the highest AAS can be seen in Table 4. The preprints which have received the highest attention score are of those which have immediate concerns, for instance medicinal interventions, and new emerging variants of COVID-19.

**Table 4:** Retracted COVID-19 Preprints.

| SL. No. | DOI | Title | Source | Reason for Retraction |
| --- | --- | --- | --- | --- |
| 1 | 10.1101/2020.01.30.927871 | Uncanny similarity of unique inserts in the 2019-nCoV spike protein to HIV-1 gp120 and Gag | bioRxiv | Concerns/Issues About Data<br>Concerns/Issues About Results<br>Notice - Limited or No Information |
| 2 | 10.1101/2021.08.10.21261836 | Efficacy and Safety of Ayurveda Intervention (AYUSH 64) as add-on therapy for patients with COVID-19 infections – An open labelled, Parallel Group, Randomized controlled clinical trial | medRxiv | Error in Analyses<br>Error in Data<br>Error in Results and/or Conclusions |
| 3 | 10.1101/2020.06.11.20128561 | Determinants of COVID-19 incidence and mortality: A cross-country analysis | medRxiv | Concerns/Issues About Data<br>Concerns/Issues About Results<br>Unreliable Results |
| 4 | 10.1101/2020.06.11.20128231 | A Model Based Analysis for COVID-19 Pandemic in India: Implications for Health Systems and Policy for Low- and Middle-Income Countries | medRxiv | Concerns/Issues about Referencing/Attributions<br>Notice - Limited or No Information |

### 10. Retracted Indian COVID-19 Preprints

Table 4 shows the Indian COVID-19 retracted preprints. There are four preprints out of 1031 which have been retracted for the reasons that have been listed in Table 5. The reasons listed in Table 4 were based on the information found on the Retraction Watch database. Two preprints were retracted because of concerns/issues about data, concerns/issue about results, and notice with very limited information on the cause of notice and unreliable results. The preprints entitled “*Efficacy and Safety of Ayurveda Intervention (AYUSH 64) as add-on therapy for patients with COVID-19 infections – An open labelled, Parallel Group, Randomized controlled clinical trial*” was retracted because of error in analysis and data. These errors in analysis and data reveal flaws in data evaluation, calculation, data collection and identification. Another preprint was retracted due to improper attribution of previous works or references, and limited information on cause of notice issued to the preprint.

## DISCUSSIONS & CONCLUSION

Preprints are the preliminary results or publications which are yet to be formally peer-reviewed by the scholarly journals. Preprints have become important piece of scholarly publications during the pandemic. There has been considerable growth in number of preprints deposited during the COVID-19 pandemic. The preprints deposited on COVID-19 related subjects have surpassed the other subject domain preprints. One out of ten preprints posted on the preprint server medRxiv in 2020 were related to COVID-19 (Else, n.d.). The COVID-19 preprints have also played an important role in disseminating and providing quick access to state-of-the-art research on COVID-19 infectious disease to find pharmaceutical interventions through vaccines and drug innovations.

The study has made an attempt to understand the Indian authors COVID-19 preprint submission practices through the bibliometric study. The studies which have explored the Indian COVID-19 related publication pattern through citation and bibliographic databases such as Web of Science, Scopus, PubMed, Dimensions and specialized WHO COVID-19 database and so on. Though dimensions.ai scholarly publication database indexes preprints, Indian COVID-19 publication related studies have hardly analyzed the COVID-19 preprints related publication pattern. This is the first study which has tried to look at the Indian COVID-19 preprints publication pattern in the bioRxiv and medRxiv preprint servers.

The study result found that, nearly 4.06% (1031 out of 25416 (18869 medRxiv preprints and 6547 bioRxiv preprints)) of the total COVID-19 preprints deposited in the bioRxiv and medRxiv preprint servers were originated from India. Previously it was found that Indians were very much reluctant to deposit their manuscript in preprint servers. Only about 3.5% Indian research papers were deposited in the arXiv preprint servers as found in the previous study (Singh et al., 2020). In a span of almost for three years there was a sizeable growth in depositing COVID-19 related preprints in bioRxiv and medRxiv preprint servers by Indian authors. This must be sustained and encourage authors to submit their research papers to preprints before submitting for scholarly journals for formal review process.

In terms posting COVID-19 preprints under various copyright and Creative Commons (CC) license it was found that though highest numbers of preprints (42.58%) were posted under CC BY-NC-ND 4.0 licenses which allows for copy and redistribute the preprint in any medium or format, 32.30% of the Indian COVID-19 preprints were posted under “All rights reserved”. This copyright license restricts others for copying or redistributing these preprints openly. This calls for creating awareness on Creative Commons License and its wider benefits for society. Preprints in the field of infectious diseases, excluding HIV/AIDS, accounted for almost 28% of the total preprints followed by epidemiology and public and global health with 22.89% and 9.89% preprints respectively. This highlights India”s significance as a source of medical information, with researchers eager to share their findings about infectious diseases on preprint servers to educate the wider community.

Another significant finding of this study is the collaborative nature of COVID-19 related preprints. Only 5.43% of the preprints were single authored preprints and remaining almost 95% of the preprints were authored by two or more authors. As this study found one-fourth of the preprints were authored by more than 11 authors. This shows the collaborative nature of COVID-19 related publications. This collaboration has allowed some authors, such as Priya Abraham (16 preprints) and Madhvi Joshi and Samiran Panda (14 preprints each), to publish a high number of preprints during the pandemic. These authors are associated with institutions such as AIIMS Delhi, IISc Bengaluru, and ICMR-NIV Pune. This highlights the contribution of Indian institutions to global medical research efforts. However, there needs to be further examination in terms of international collaborative pattern of authors of Indian COVID-19 preprints. The authors such as Priya Abraham, Nivideta Gupta, Pragya Yadav, others and the major institutions such as AIIMS, New Delhi, Rishikesh, ICMR, New Delhi and ICMR-NIV, Pune associated with highest number of preprints to their credits have also published highest number of peer-reviewed research papers as well (Pathak, 2020; Vasantha Raju & Patil, 2020). The authors with highest number of COVID-19 preprints have all come from same institutions (e.g., ICMR-NIV, Pune) or associated with their parent institution (e.g., ICMR-New Delhi). There is a more intra-institutional collaborative research than international collaboration on COVID-19 infectious disease.

As many as 283 preprints out of 1031 COVID-19 preprints have published in peer-reviewed journals. This accounts for 27.45% of the total preprints deposited in bioRxiv and medRxiv. One significant findings of the study is that 49.17% (118 out of 240 preprints) of the Indian COVID-19 preprints deposited in bioRxiv have published in journals as compared to medRxiv preprints where 20.86% of preprints have published in peer-reviewed journals. In a study done by (Lachapelle, 2020) has found that 19.6% of the COVID-19 preprints deposited in arXiv, bioRxiv and medRxiv preprint servers between January and early September, 2020 have published in peer-reviewed journals. (Abdill & Blekhman (2019) have found that 75% of the preprints deposited in bioRxiv preprints have published in peer-reviewed journals within eight months period. More or less our study concur the findings reported in the study done by Lachapelle (2020) and Abdill & Blekhman (2019). However, we have observed that there were few preprints which have appeared in journals but have not been reflected in the NIH iSearch COVID-19 portfolio database.

Another interesting finding of the study is that in total 173 journals have published 283 COVID-9 preprints. Of this 173 journals, PLOS One has published highest number of preprints (14 out of 283), followed by Scientific Reports with 10 preprints published in its journals. Other top journals that published COVID-19 related preprints includes Infection, Genetics and Evolution, International Journal of Infectious Diseases, Frontiers in Immunology, Journal of Family Medicine and Primary Care, Virus Research and others. Most of these journals have good impact factors and high journal quartile ranking. To name few-Journal of Infection (IF 38.637), Clinical Infectious Diseases (IF 20.999), Journal of Medical Virology (IF 20.693), International Journal of Infectious Diseases (IF 20.999), eBioMedicine (IF 11.205), Science of The Total Environment (10.754) and others. Studies have also reported that posting manuscripts in preprint servers contributes to the acceptance of papers in peer-reviewed journals (Lin et al., 2020). With elife Sciences (eLife Sciences Publications Ltd, 2023) recent announcement of publishing all peer-reviewed manuscripts as Reviewed Preprints and do away with accept/reject decisions of submitted manuscripts will be radically alter how scientific publication being communicated so far. With this kind of radical change in the scholarly publishing landscape preprints will be accepted by the larger scientific community.

One of the significant advantages of depositing manuscript in preprint server is the chances of increasing number of citations and online mentions in quick time (Fraser et al., 2020). In this study also it was observed that in quick time many COVID-19 preprints have received considerable number of citations. For instance, “Repurposed antiviral drugs for COVID-19-interim WHO SOLIDARITY trial results” has received 263 citations, and another preprint “Convergent evolution of SARS-CoV-2 spike mutations, L452R, E484Q and P681R, in the second wave of COVID-19 in Maharashtra, India” has received 227 citations. The top 10 Indian COVID-19 preprints that we have listed have received more than 100 citations each. Previous studies have also showed that preprints which were published subsequently in peer-reviewed journals have received more citations and online mentions compared to papers directly submitted to the journals and subsequently published after due peer-review process (Davis & Fromerth, 2006; Serghiou & Ioannidis, 2018). There were four COVID-19 Indian preprints retracted for various reasons mainly issues with data. One of the preprint which was widely discussed and critiqued on online social networking platforms on claiming to found similarities between the new coronavirus and HIV was retracted in quick time and it was appreciated by the scientific community for its swiftness in which this preprint was taken down (Oransky & Marcus, 2020). A study found that preprints helps in faster identification of erroneous publications as compared to traditional peer-reviewed journals (M. Singh et al., 2022)

Submitting or depositing manuscripts in preprint servers has a greater benefit such as instant or rapid peer feedback, wider reach and visibility, and possible collaborative research opportunities, etc. This pandemic has shown how preprints have played an important role in providing quick and open access to research results for mitigating the spread of COVID-19 virus. As this study showed that Indian authors have also embraced preprint servers to deposit their COVID-19 manuscripts in large numbers during this pandemic. This needs to be sustained and those authors who deposit their preprints should be incentivized by institutions to encourage them to make their publications and data open and reap wider benefits of open science.

## LIMITATIONS

This study has examined only COVID-19 preprints deposited in bioRxiv and medRxiv preprint servers, and these preprint servers accept preprints largely from biomedical sciences. Our study dominantly focused on COVID-19 preprints submitted on biomedical sciences. The NIH *iSearch* COVID-19 Portfolio database includes COVID-19 preprints deposited in five other preprint servers mainly arXiv, preprints.org, Research Square and others. The Research Square preprint server is also highly populated with COVID-19 preprints. Future studies can include Indian COVID-19 preprints submitted to arXiv, ChemRxiv, Preprints.org, Research Square, and Qeios to understand the comprehensive overview of preprinting practice of Indian authors during the pandemic. The survey research can also be combined with the quantitative approach to elicit the opinion of Indian authors on what made them to deposit their COVID-19 manuscript in preprint servers to better under this phenomenon. The author affiliation data was manually collected for this study, thus few omissions in the data may not be ruled out.

## CONCLUSION

The COVID-19 pandemic has elicited a pressing need to quickly communicate research findings to the scholarly community. To meet this need, preprint servers have become an increasingly important platform for sharing preliminary research findings. These servers provide a way to disseminate insights and directions for future research. This is in line with UNESCO”s mission to promote open science and the collaboration between publishers and stakeholders to make research accessible to everyone. The study found that Indian researchers have embraced preprint servers, such as bioRxiv and medRxiv, as a means of rapidly sharing their findings with the wider community. This is particularly important in the fight against the pandemic, where swift and effective communication of research is essential. The study concludes that the active participation of Indian researchers in using preprint servers underscores the significance of open science in addressing critical global issues.

## Data Availability

Data can be obtained upon request from the Corresponding Author

## Supplementary File-1

**https://drive.google.com/file/d/1Zh017vYTHIwGCVg4qxFKCJYSLHAO5sRZ/view?usp=share_link**

## Acknowledgment

We extend our sincere gratitude to Amrollah Shamsi, an insightful Independent Researcher based in Bushehr, Iran, for his invaluable contributions and enlightening discussions throughout the development of this project.

## Notes

**Conflict of Interest:** The authors have no financial or proprietary interests in any material discussed in this manuscript.

### Competing Interest Statement

The authors have no financial or proprietary interests in any material discussed in this manuscript.

### Funding Statement

This Study received no specific grant from any funding agency

