## Supplementary File-1 for "Indian COVID-19 Preprints Submissions in bioRxiv and medRxiv Preprint Servers"

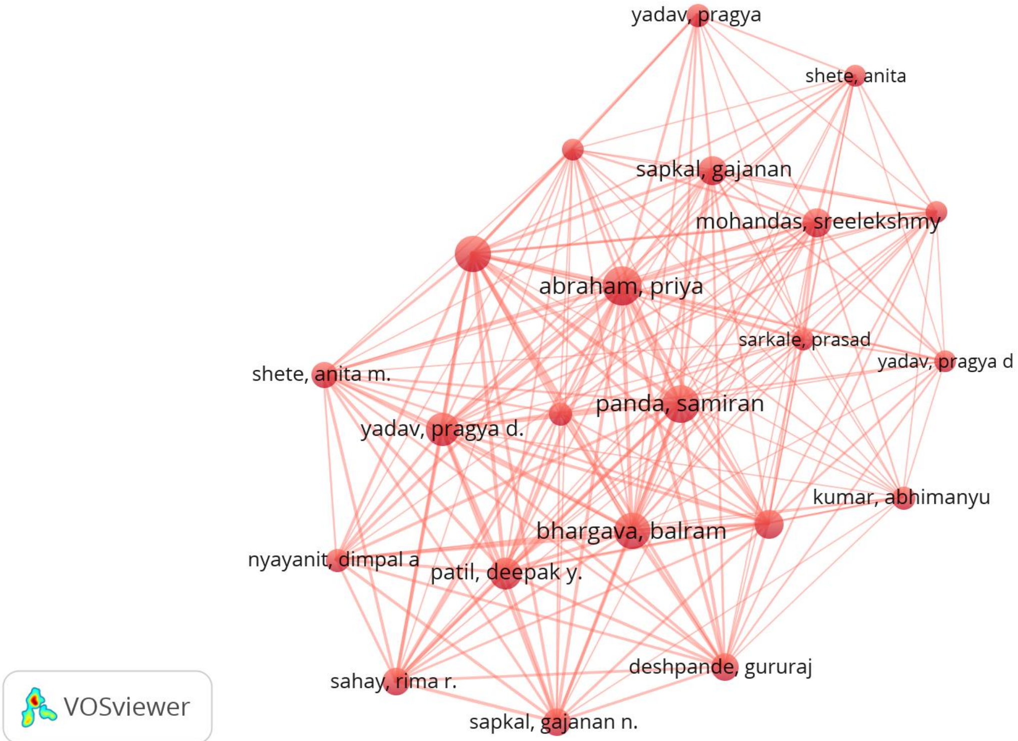


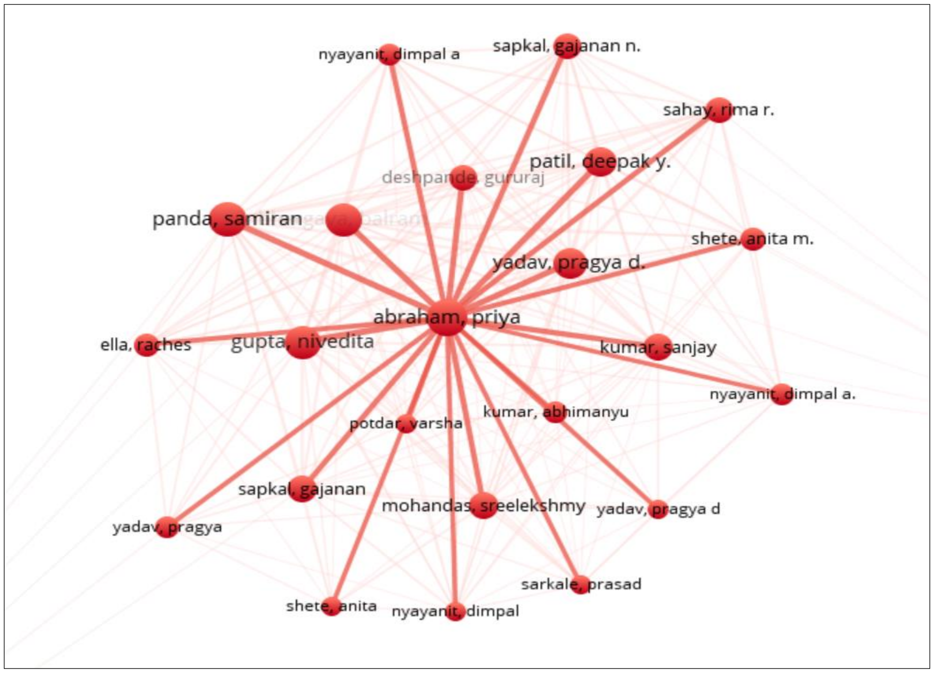


**Figure-S1: Collaborative Authorship Network Visualization of Priya Abraham of ICMR-NIV, Pune**
